# Epidemic dynamics in inhomogeneous populations and the role of superspreaders

**DOI:** 10.1101/2021.02.08.21251386

**Authors:** K. Kawagoe, M. Rychnovsky, S. Chang, G. Huber, L. M. Li, J. Miller, R. Pnini, B. Veytsman, D. Yllanes

**Author notes:** These authors contributed equally to this work (K. Kawagoe, M. Rychnovsky).

## Abstract

A variant of the SIR model for an inhomogeneous population is introduced in order to account for the effect of variability in susceptibility and infectiousness across a population. An initial formulation of this dynamics leads to infinitely many differential equations. Our model, however, can be reduced to a single first-order one-dimensional differential equation. Using this approach, we provide quantitative solutions for different distributions. In particular, we use GPS data from ∼ 10^7^ cellphones to determine an empirical distribution of the number of individual contacts and use this to infer a possible distribution of susceptibility and infectivity. We quantify the effect of superspreaders on the early growth rate ℛ_0_ of the infection and on the final epidemic size, the total number of people who are ever infected. We discuss the features of the distribution that contribute most to the dynamics of the infection.

## I. INTRODUCTION

A strong temptation in modeling a system consisting of many similar parts is to make the assumption that these parts have identical properties. Accordingly, the classical models in epidemiology assume (often implicitly) that everyone has the same propensity to be infected and, if infected, the same propensity to infect others [1]. This assumption may be justified when differences in the salient parameters are small. However, one of the interesting features of the current COVID-19 pandemic is the huge variation in infectivity: small numbers of infectious events or individuals seem to be responsible for a large number of cases [2–14]. This feature seems to be present in other coronavirus epidemics including SARS [15–17] and MERS [18–20]. One can point to different explanations for this phenomenon: individual variations in viral load and shedding [14, 21, 22], in droplet production (see the review in [23]), in contact networks [8, 10, 12, 24, 25], and differences in the features of ventilation systems at certain events and venues [26, 27]. Inhomogeneity seems to have played an important role for other epidemics as well [28–30], leading to the rule of thumb that “20% of patients produce 80% of infections” [31]. However, it seems that for coronavirus-related infections the variability is even higher than that heuristic [13, 14]. In a recent book [32], the historian Lepore has noted that “the study of the human condition is not the same as the study of the spread of viruses and the density of clouds and the movement of the stars,” which is incontrovertible. The converse, however, is not: it appears that the spread of viruses *is* dependent on at least one aspect of the human condition, namely the intrinsic variability and lack of uniformity of human behavior.

There are two related, but distinct, notions of superspreading in this literature, namely, *superspreading events* and *superspreading individuals*. Superspreading events are events that produce many infections. Super-spreading individuals (*superspreaders*) are specific people that produce many infections (such as Typhoid Mary in the early 1900s). As one might imagine, in reality, some combination of these two processes is present. In this paper, however, we set our sights on the latter phenomenon: a superspreader is always an individual, rather than an event.

It is reasonable to assume that a variability in infectivity is accompanied by a variability in susceptibility. Common explanations of variability in individual infectivity — increased shedding due to higher rate of virus multiplication in the given host, increased exposure period, and increased personal contacts — suggest that increased infectivity may correlate with increased susceptibility. We note that there are arguments for the opposite correlation: for example, some studies indicate that older age may correspond to higher susceptibility but lower infectivity [33], while other studies seem to contradict this finding [34]. The change of contact patterns caused by mitigation measures further confounds this issue [35]. However, the assumption of positive correlation between infectivity and susceptibility seems to be a reasonable one. One conclusion of this is that superspreaders might be more prominent at the early stages of an epidemic. During the course of an epidemic, the fraction of super-spreaders will typically decrease with time. This would lead to a change in the apparent value of the average transmission rate, which could make it difficult to evaluate the effectiveness of mitigation measures. This effect might be quite large and is not captured by many standard models. Generally speaking, inhomogeneity may lead to a large change in the mean behavior of a system, especially when fast growth is involved. Understanding the effect of inhomogeneity would increase the fidelity of models based on real-world data, and lead to more effective public policy.

Several recent works (see, e.g., [9–12, 25, 36–38]) have addressed the issue of heterogeneity in the population, but they either concentrate on specific distributions or treat the variability in infectivity and susceptibility separately, without considering the effect of a possible correlation between the two. Ref. [10] in particular reports numerical experiments with heterogeneous quenched contact probabilities (ours are uniform). A rich set of outcomes was obtained that calls for an analytical treatment in the flavor of this manuscript.

In this work we discuss the epidemic dynamics for a population with variable infectivity potential accompanied by variable individual susceptibility. We obtain the results for the general case of an arbitrary distribution of susceptibility and infectivity. We also provide a calculation of ℛ_0_ that quantifies the unintuitive effect of superspreaders on the early growth rate of the epidemic and find that it depends strongly on the correlation between susceptibility and infectivity.

Furthermore, one of the distributions holds a special interest. If we assume that the main driver of inhomogeneity is diversity in the number of social contacts for an individual, then data [39] on the distribution of these contacts suggests a very wide distribution of infectivity and susceptibility.

An important question for modeling the inhomogeneity is whether the result depends only on the moments of the distribution (mean, variance, skewness, …) or on the behavior of the tails of the distribution. The answer to this question could inform the construction of realistic predictive models in the future. We discuss both the cases of fat tails and skinny tails, and the transition between these regimes.

We recently became aware of work by Tkachenko and collaborators [40] that also employs a model with distributed infectivity and susceptibility of the population. The treatments of the issue in [40] and the present paper are, however, different. We attempt to study the range of effects caused by a varying degree of correlation in a systematic way, in the framework of a minimal model. Tkachenko et al., on the other hand, postulate a one-to-one correspondence between susceptibility and infectivity. This assumption is a limiting case for our model, corresponding to the “worst-case scenario” for the epidemic, as we show in Appendix C. In terms of fits to real-world data, Tkachenko et al. consider time series of hospitalizations in Chicago and NYC to calibrate their model, while we take a different approach and work from anonymized GPS data from cell phones to infer empirical distributions of the number of social contacts. In situations with matching assumptions, the conclusions of [40] and the present paper are in agreement. We believe that, together, these two works present a nicely complementary view of correlation between susceptibility and infectivity in epidemics.

The rest of the paper is organized as follows: In Section II, we give a mathematical description of the dynamics of our model. In Section III, we reduce our model to a one dimensional integro-differential equation, analyze the long time dynamics, and describe an early time criterion for epidemic outbreak. In Section IV, we compare the results of our model for different distributions of population attributes, including an empirical one from anonymized cell phone data. We end with our discussion and conclusions in Section V. In the Appendices, we provide derivations which are relevant to the main text and we discuss some of the methodological aspects of our empirical data.

## II. THE MODEL

Classic SIR models [1] divide the population into three compartments: susceptible *S*, infected *I*, and recovered (or deceased) *R*. The rate of new infections in this model is proportional to the number of encounters of susceptible persons with the infected persons, while the rate of recovery is proportional to the number of infected persons. This gives us the well-known SIR equations

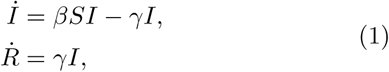

where *S, I*, and *R* are the fractions of susceptible, infected, and recovered persons to the constant population size, dot means the time derivative, *β* and *γ* are non-negative constants, and we use the fact that, with our normalization, the fraction of susceptible persons *S* satisfies the equation

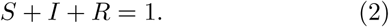

We use the simplest version of the model, which accounts neither for additional births and deaths, nor for population migration. Additionally, we do not allow for the possibility of recovered individuals being reinfected.

We now allow the parameters to be different for different individuals. Namely, let the infection rate *β* in equation (1) be the product of individual susceptibility *s* and infectivity *σ*. To obtain the rate of infection, we integrate over the values of *s* for susceptible individuals and over the values of *σ* for infected individuals. Note that in our model the values of *s, σ*, and *γ* are fixed for each person and do not change with time.

Let *p*(*σ, s, γ*) d*σ* d*s* d*γ* be the probability that a person selected uniformly at random from the population has susceptibility *s*, and, when infected, has infectivity *σ* and recovery rate *γ*. Note that *p* does not change with time in our model. We will have reason to make repeated use of the averaging operator 𝔼: for any function *f* (*σ, s, γ*), we define

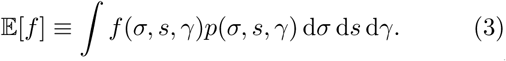

Equations (1) and (2) should now be rewritten, because *I, R* and *S* are not just functions of time *t*, but also depend on *s, σ*, and *γ*. Namely, let *I*(*σ, s, γ, t*) *dσ ds dγ* be the probability that a person selected uniformly from the entire population at time *t* is infected and has (initial) susceptibility *s*, infectivity *σ* and recovery rate *γ*. Similarly we introduce *S*(*σ, s, γ, t*) and *R*(*σ, s, γ, t*). Then equation (2) becomes

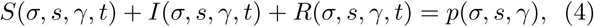

and equations (1) become

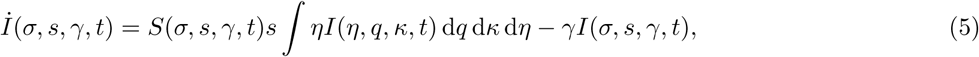

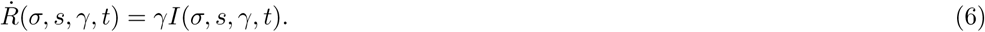

When the proportion of infected individuals is small, *S*(*σ, s, γ, t*) in equation (5) is close to *p*(*σ, s, γ*), giving a linear approximation of equation (5). For distributions where is *γ* a constant, it can be shown (Appendix B) that the early behavior of an epidemic is determined by ℛ_0_ = 𝔼[*σs*]/*γ*.

The total fraction *Ω*(*t*) of persons who have ever been infected at time *t* is the sum of currently infected and recovered individuals. If we stratify *Ω* by *s, σ*, and *γ*, we can write down

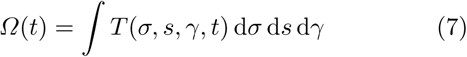

with

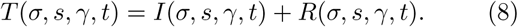

The final epidemic size is

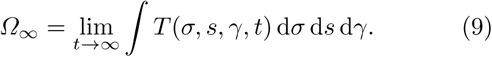

We will use index 0 for the initial conditions in equations (5) and (6), so *I*_0_(*σ, s, γ*) = *I*(*σ, s, γ*, 0) etc.

## III. ANALYTIC RESULTS

In this section we discuss the general properties of our model. We assume that the distribution of infectivity and susceptibility is such that the moments 𝔼[*σ*], 𝔼[*s*], and 𝔼[*σs*] as defined in equation (3) exist. If the distribution is so heavy tailed that these moments do not exist then important integrals in our analysis will not converge. This is not merely a technical restriction. For instance the short time behavior of the model should be quite different if 𝔼[*σs*] is infinite.

Let us introduce the notation:

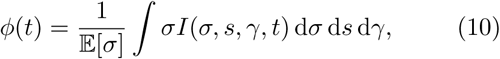

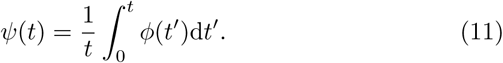

An individual has infectivity *σ* if infected and 0 if not. Therefore, 𝔼[*σ*] is the maximal average infectivity (when everyone is infected simultaneously), and *ϕ*(*t*) is the ratio of the current average infectivity and the maximal one. Further, *ψ*(*t*) is the historical average of *ϕ*(*t*). Both these quantities are thus between zero and one. In our model (without births or immigration and no persons with zero recovery rate), there are no infected persons at *t* → ∞, so in this limit

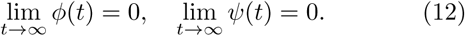

It is shown in Appendix A that the stratified fraction of people who ever have been infected at time *t* [see equations (7) and (8)] is

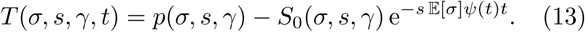

For outbreaks started with a small number of infected persons, almost all remaining individuals are susceptible, so *S*_0_ ≈ *p*. The number of currently infected individuals is

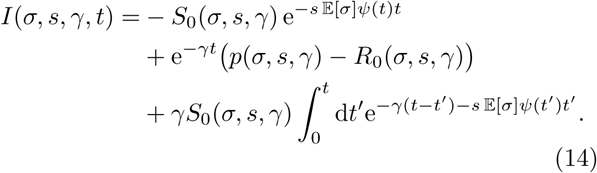

As a result, if we know *ψ*(*t*), then we know the full solution. It is shown in Appendix A that *ψ*(*t*) is a solution of the equation

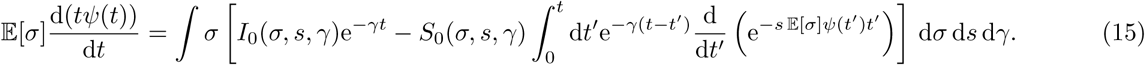

To study the behavior of equation (15) we will make several simplifying assumptions. First, we assume a constant recovery rate across the population:

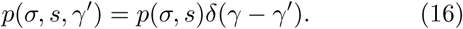

This means that the other variables (*S, I, R*) are also proportional to *δ*(*γ* − *γ′*); we will use the same notation for them as functions of *σ* and *s*.

Second, we assume the initial number of recovered individuals is zero,

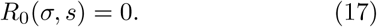

Third, we assume that the initial distribution of infected persons is proportional to *p*(*σ, s*), and is small:

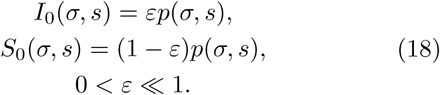

To see why any other initial distribution *I*_0_ that is small should behave similarly see Appendix B.

With these assumptions equation (15) can be further transformed from an integro-differential equation to a first-order differential equation

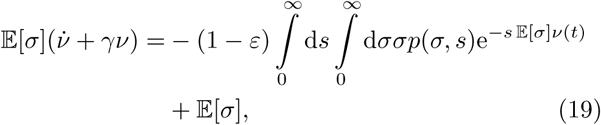

for the function *ν*(*t*) = *ψ*(*t*)*t* (See equation (A14)).

To numerically solve equation (15) it is convenient to rewrite it as two first-order differential equations (See Appendix E). In the rest of this section we discuss the properties of the solution of this equation.

Let us start with the final epidemic size [equation (9)]. It can be shown (Appendix A) that at *t*→ ∞ the function *ψ*(*t*) in equation (11) behaves as 1/*t*. Choose *L* so that at large *t*,

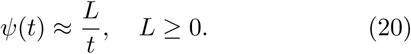

Then equation (9) with *T* from equation (13) becomes (see Appendix A)

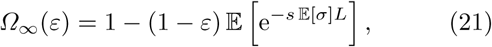

where *L* is the unique nonnegative root of the equation

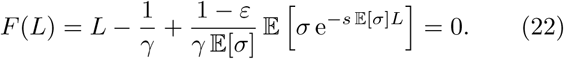

We are interested in an infection started with a small number of initial cases, which corresponds to *ε* → 0. If in this limit equation (22) has a strictly positive root, the final epidemic size

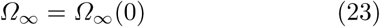

is non-zero, and does not depend on *ε*: in other words, the epidemic takes off. If the limit does not have a strictly positive root then the infection immediately dies out and the final epidemic size is 0. In this *ε* → 0 limit *F* (0) = 0 and *F* (1/*γ*) *>* 0, so equation (22) has a positive (non-zero) root if d*F* (0)/d*L <* 0. Taking the derivative, we see that a non-zero root corresponds to the condition

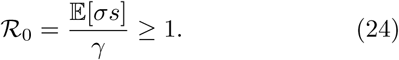

Given this result, we take a brief detour from our discussion of *t*→ ∞. Another way to look at epidemic spread is to study the short term behavior of the solution. Our analysis (Appendix B) shows that the initial small infection spreads with exponential rate ℛ_0_ = 𝔼[*σs*]/*γ* determined by equation (24). The upshot is that the growth rate of the epidemic is highly dependent on how correlated the infectivity and susceptibility are.

One naive generalization of ℛ_0_ from the SIR model, i.e., the average number of secondary infections produced by a typical infection would be 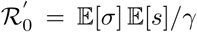. To explain why ℛ_0_, rather than 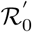, determines the exponential growth rate of the infected population we will illustrate what the two quantities measure. If we choose a person from the *entire* population uniformly at random and infect them, then the average number of secondary infections would be 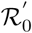. For instance if a cruise ship travels somewhere and almost everyone is infected, then when they return home the expected number of secondary infections each person produces will be 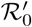. On the other hand a person who was infected via community spread (early in the epidemic) will cause on average ℛ_0_ secondary infections. The difference between these cases is that in the first case almost all travelers are infected so the fact that someone is infected tells us little about their susceptibility, whereas in the second case people are infected via community spread which occurs with a probability proportional to their susceptibility early in the epidemic. See Appendix B for details.

We will now continue our discussion of the final epidemic size with some limiting cases. As mentioned above, for an epidemic to spread, it is necessary that ℛ_0_ = 𝔼[*σs*]/*γ* ≥ 1. Near this transition, where ℛ_0_ ≈ 1, we may write down an approximation for *L*. Again, we will be interested in the limit of small initial epidemic size *ε* → 0, although it is not difficult to generalize the following result for non-zero *ε*. Let ℛ_0_ *>* 1. Assuming that *L* is small, and that *p*(*σ, s*) falls off quickly enough for large *s*, we may approximate equation (22) as

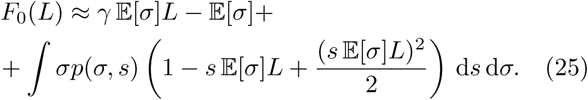

Therefore, if we get close enough to the transition where 𝔼[*σs*] − *γ* is small

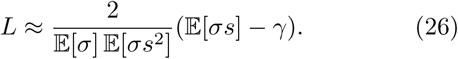

In this regime, equation (21) gives the total epidemic size as

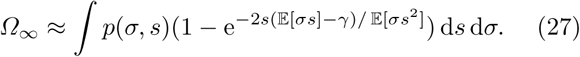

Let us now briefly discuss the opposite limit. Instead of *γ* being so large that the epidemic almost doesn’t start, we study *γ* so small that the epidemic infects almost everyone. It is expected that if *γ* = 0, then the entire population will eventually become infected; that is, *Ω*_∞_ = 1. Equation (21) shows that in this case *L* → ∞. It is easy to show that for small *γ, L* ≈ 1/*γ*, and equation (21) predicts an exponentially small number of individuals not infected.

This framework allows one to make predictions for a number of specific distributions discussed in the next section. We conclude the general discussion with one very interesting case: when the distribution has a very small number of “superspreaders”, individuals with anomalously high infectivity. (Here very small means small enough to not appreciably change 𝔼[*σs*].) A relevant question is whether these individuals have an oversized contribution in the epidemic. Equations (21) and (22) show that this is *not* the case, and the contribution of superspreaders is limited by the linear term in the average value of 𝔼[*σs*] (see Appendix D). Therefore, while superspreaders still contribute to the dynamics, they are only a primary driver of infection in our model when they significantly change ℛ_0_ if their number is large.

## IV. RESULTS FOR DIFFERENT DISTRIBUTIONS OF INFECTIVITY AND SUSCEPTIBILITY

Let us further illustrate the general results using specific distributions for *s* and *σ*. First, consider an *N* - component SIR model. That is, there are *N* different types of individuals who have parameters *σ*_*i*_, *s*_*i*_, *γ*_*i*_ and represent a portion of the population *p*_*i*_, and

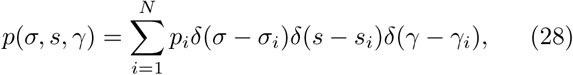

*δ*(*x*) being Dirac’s delta-function. In the case where *N* = 1, this reduces to the standard SIR model. We see in Appendix B that this model is a limiting case of the model presented in this paper [41].

Another useful distribution to study is the Gamma distribution with *σ* = *s*. In particular, we are interested in the distribution

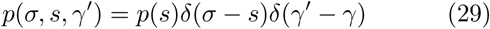

where

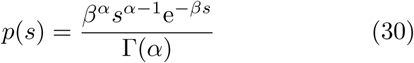

and *α, β* are positive constants. This system is interesting to study because the integrals involved in solving for *L* are analytically tractable. In the case where *α* = 1 we recover the exponential distribution and we can find *Ω*_∞_ exactly (equation (F6)). We analyze the case of the Gamma distribution in Appendix F.

We further illustrate the dynamics of epidemics using several special cases of distributions of infectivity *σ* and susceptibility *s* with the assumption of constant recovery rate *γ*. (See Appendix C for an analysis of which distributions lead to the worst outcomes for the final epidemic size.)

Even with constant *γ* the answer depends on the probability distribution *p*(*σ, s*). We discuss three limiting cases: (i) completely independent *σ* and *s*, with *p*(*σ, s*) = *p*_*σ*_(*σ*)*p*_*s*_(*s*); (ii) completely positively correlated *σ* and *s* with *σ* ∝ *s*; and (iii) positively correlated *σ* and *s* with a correlation coefficient *ρ*.

Note that since only the product *σs* enters the equations, we always can multiply *σ* by a constant factor *f*, and *s* by the factor 1/*f*. We choose this factor to ensure that 𝔼[*σ*] = 𝔼[*s*]. In the numerical calculations in this section we used the following parameters roughly following [42–44]

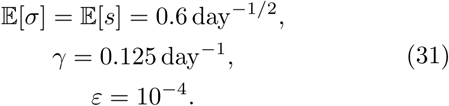

At present, our understanding of variability in individual susceptibility and infectivity is far from complete. While the consensus is that they have a wide distribution (see the discussion in the Introduction), the shape of this distribution is not known, and most studies assume a convenient one for their calculations. Since we want to explore the dependence of the dynamics on the distribution itself, rather than on its parameters, we compare two reasonable *a priori* assumptions: a log-normal distribution with the parameters *µ* and 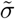, and a Gamma distribution with the parameters *α* and *β*. Another approach is to suggest some mechanism for the variability and choose a distribution that follows this mechanism. One such mechanism is the variability of individual contacts: the more contacts has a person, the higher is their *s* and *σ*. It is important to note that in this model *s* is completely correlated with *σ* because they are caused by the same mechanism.

We are fortunate to be able to use empirical data about the number of contacts from the “path-crossing” network described in Looi et al. [39]. Their network is constructed from the mobility data provided by SafeGraph, a company that aggregates and anonymizes geolocation data from cell phone applications. SafeGraph collects GPS location pings for millions of adult smartphone users in the United States, where each ping represents the latitude and longitude of one user at one timestamp. Looi et al. [39] transform the set of location pings into a dynamic network, where users are represented as nodes, and edges indicate the number of times two users crossed their paths (see Appendix G for the details). We use the number of path crossings as a proxy for the number of users’ social contacts, which is in its turn a proxy for susceptibility and infectivity. Due to the number of assumptions here one should be careful with the interpretation of the results. We do not claim that the SafeGraph data provide *the* distribution of *σ* and *s*. Rather we think they suggest features of the real distribution. Moreover, it should be stressed that this is just one of many possible mechanisms for susceptibility and infectivity heterogeneity, see, e.g., the discussion in [13]. We do not claim that this is the only, or even the main mechanism —it is just the one for which we have data.

An interesting feature of the SafeGraph distribution is that it is very wide. The average number of contacts per user is 0.342 × 10^3^, while the standard deviation is 1.04 × 10^3^. We can try to approximate the empirical distribution of contacts using a theoretical distribution. On Figure 1 we show log-normal and gamma approximations together with the empirical distribution with the same mean and variance.

**Figure 1.**
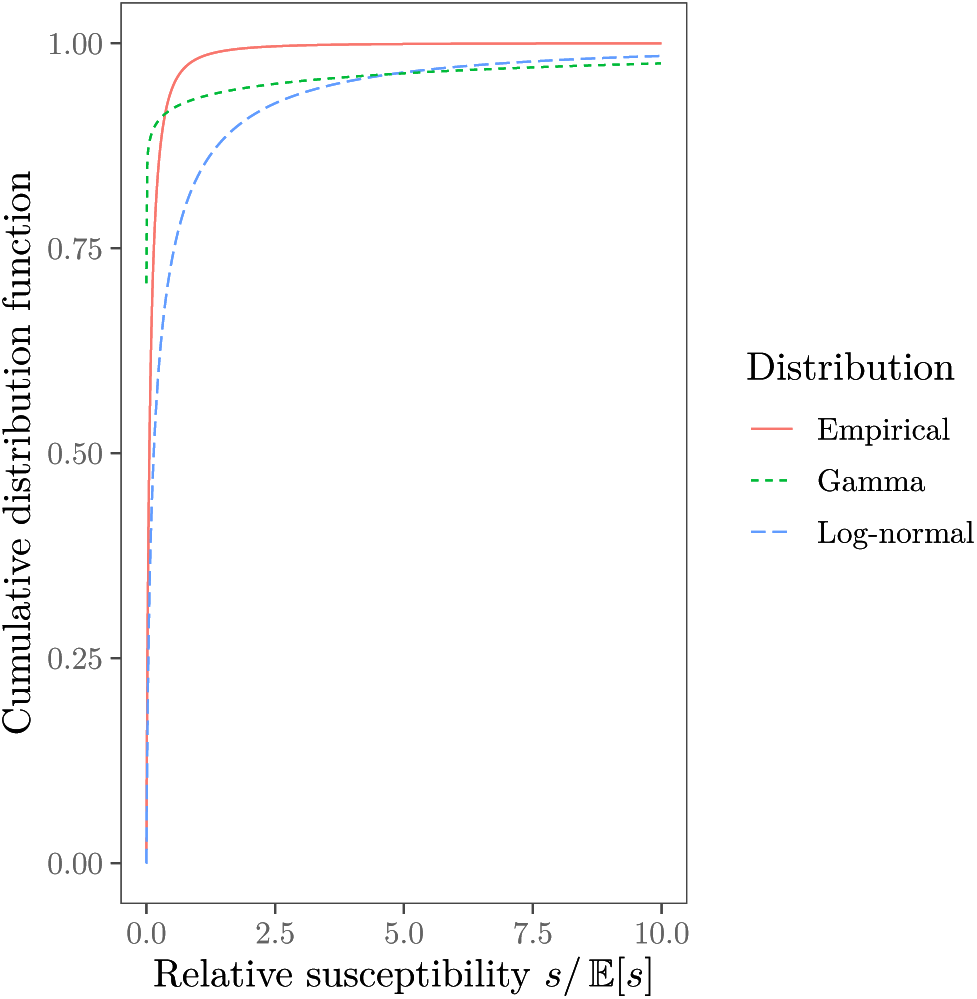
Comparison of empirical, log-normal and gamma distributions with the same average infectivity 𝔼[*s*] = 0.6 day^−1/2^ and variance *ζ*^2^ with *ζ* = 4.16 day^−1/2^.

In the remainder of this section we discuss the numerical solutions of the model equations for the log-normal, Gamma, and empirical distributions obtained with the approach discussed in Appendix E. See Appendix F for analytical solutions in special cases.

In Figure 2 we compare the epidemic’s progression for log-normal and Gamma distributions with the same mean *s* and varying distribution widths. We see that a wider distribution leads to a lower epidemic size. When the width of the distribution decreases, the curve goes to the one for the classical SIR model. An interesting feature is that a wide correlated distribution of *s* and *σ* leads to an earlier start of the epidemics instead of the S-like curve of the standard SIR model.

**Figure 2.**
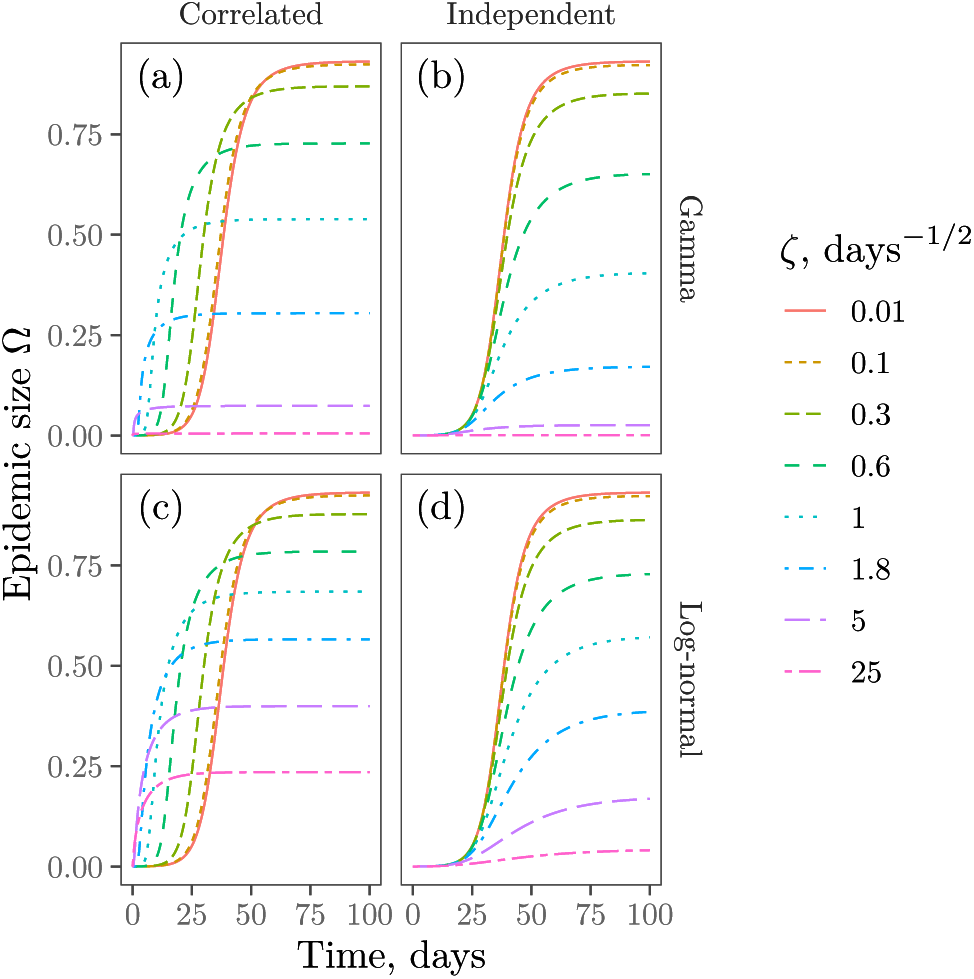
Comparison of epidemics spread for Gamma [panels (a)–(b)] and log-normal [panels (c)–(d)] distributions of infectivity and susceptibility with standard deviation *ζ* and parameters in equation (31). The cases of independent or completely correlated *σ* and *s* are shown.

In Figure 3 we study the influence of the positive correlation between infectivity and susceptibility. For simplicity we show just the final size *Ω*_∞_. As demonstrated by this figure, the more correlated these parameters are, the higher the size is, as predicted by the analysis in the previous section.

**Figure 3.**
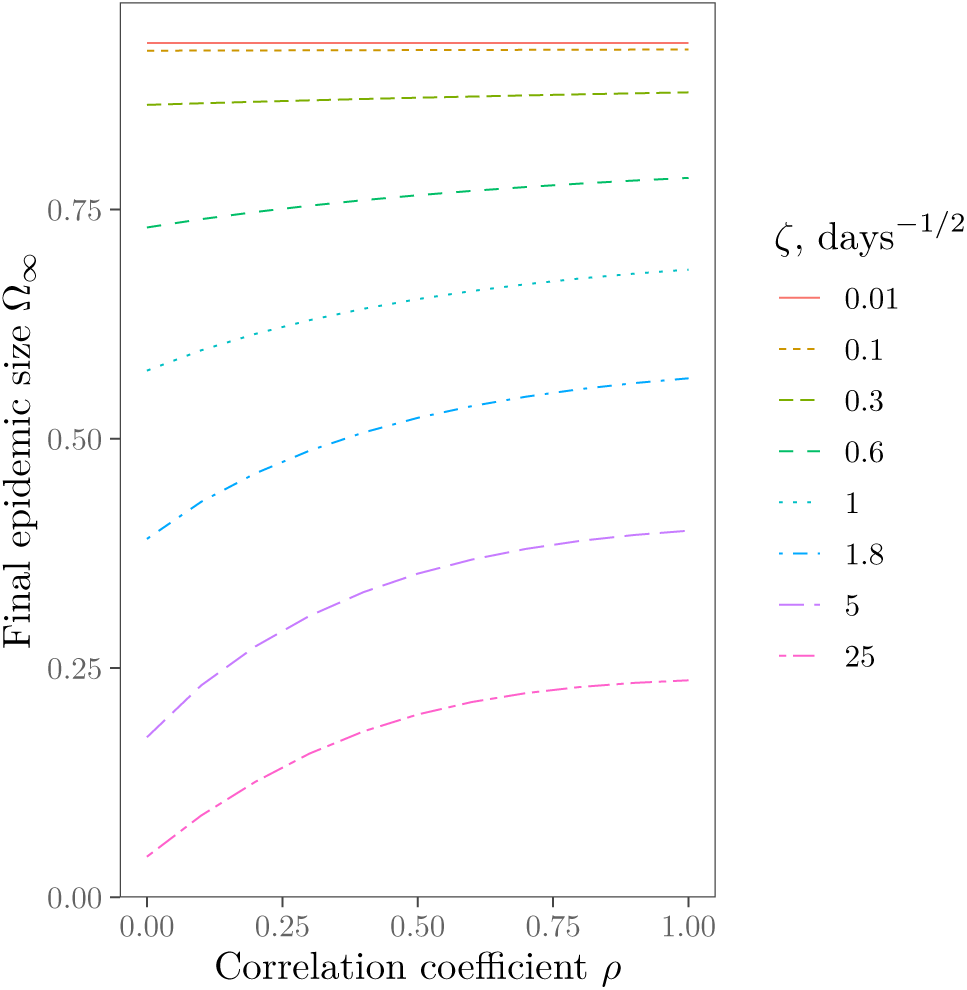
Dependence of final epidemic size *Ω*_∞_ on *ρ* where [ln(*s*), ln(*σ*)] is a Gaussian vector with mean 𝔼[*s*] = 𝔼[*σ*] = 0.6 day^−1/2^, Var(*s*) = Var(*σ*) = *ζ*^2^ and correlation 𝔼[*sσ*] = 𝔼[*s*] 𝔼[*σ*] exp(*ρς*^2^) with *ς*^2^ ≡ ln[1 + *ζ*^2^/(𝔼[*s*] 𝔼[*σ*])]. Note that *ρ* is the correlation coefficient for ln(*s*) and ln(*σ*) rather than for *s* and *σ*.

For another comparison we take the empirical number of contacts between the individuals (Appendix G) as a proxy for both *s* and *σ*. We renormalize the number of contacts to obtain the average infectivity 𝔼[*s*] in equation (31). This leads to variance *ζ*^2^ = 17.27 day^−1^ (*ζ* = 4.16 day^−1/2^). Then we fit the parameters of log-normal and Gamma distributions to get the same 𝔼[*s*] and *ζ*. All three distributions are shown on Figure 1.

The results are shown in Figure 4 together with the solution for the classical SIR model with the infectivity and susceptibility equal to the averages 𝔼[*s*] and 𝔼[*σ*].

**Figure 4.**
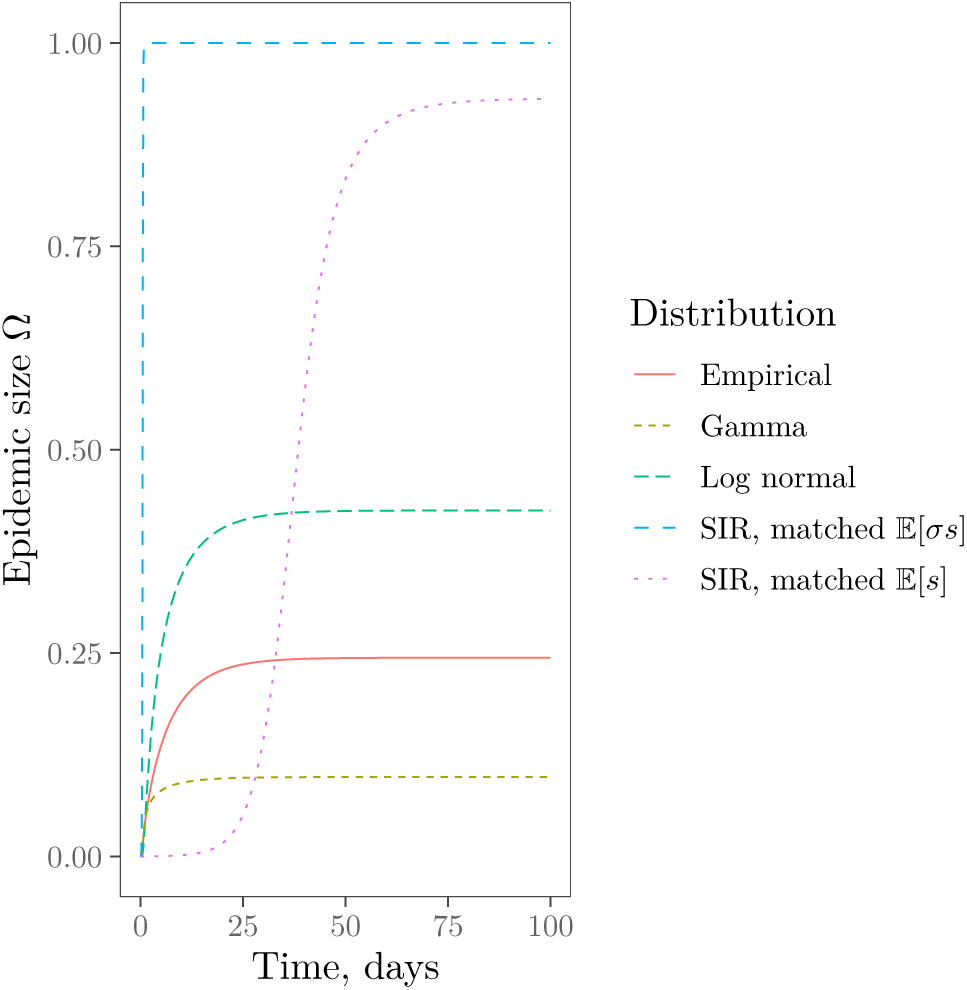
Epidemics progression for the distributions shown on Figure 1 with parameters in equation (31). A classical SIR solution for the same susceptibility and infectivity is also shown.

The figures suggest that, generally speaking, variability in susceptibility and infectivity lowers the final epidemic size, and the correlation between them increases it. Important special cases of this statement are proven in Appendix C, and based on the figures, we expect it to hold more generally.

Of special interest is the question of whether individuals with high infectivity (“superspreaders”) influence the epidemic dynamics and final epidemic size. To model the effect of superspreaders we can discuss a special bimodal distribution of infectivity,

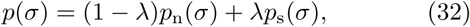

where *p*_n_ describes “normal” persons with low *σ*, and *p*_s_ describes superspreaders with high *σ*. In our numerical experiments we modeled superspreaders using a powerlaw distribution

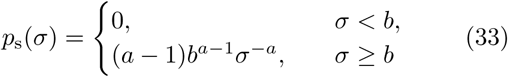

with the parameters *a* = 4, *b* = 1.2 day^−1/2^. With these parameters the average infectivity of superspreaders is 1.8 day^−1/2^, i.e., three times the average infectivity in our simulations. The results are shown on Figure 5. We see that the influence of superspreaders is at most linear in their proportion *λ*. This is not coincidental: as shown in Appendix D, the effect of superspreaders is at most linear.

**Figure 5.**
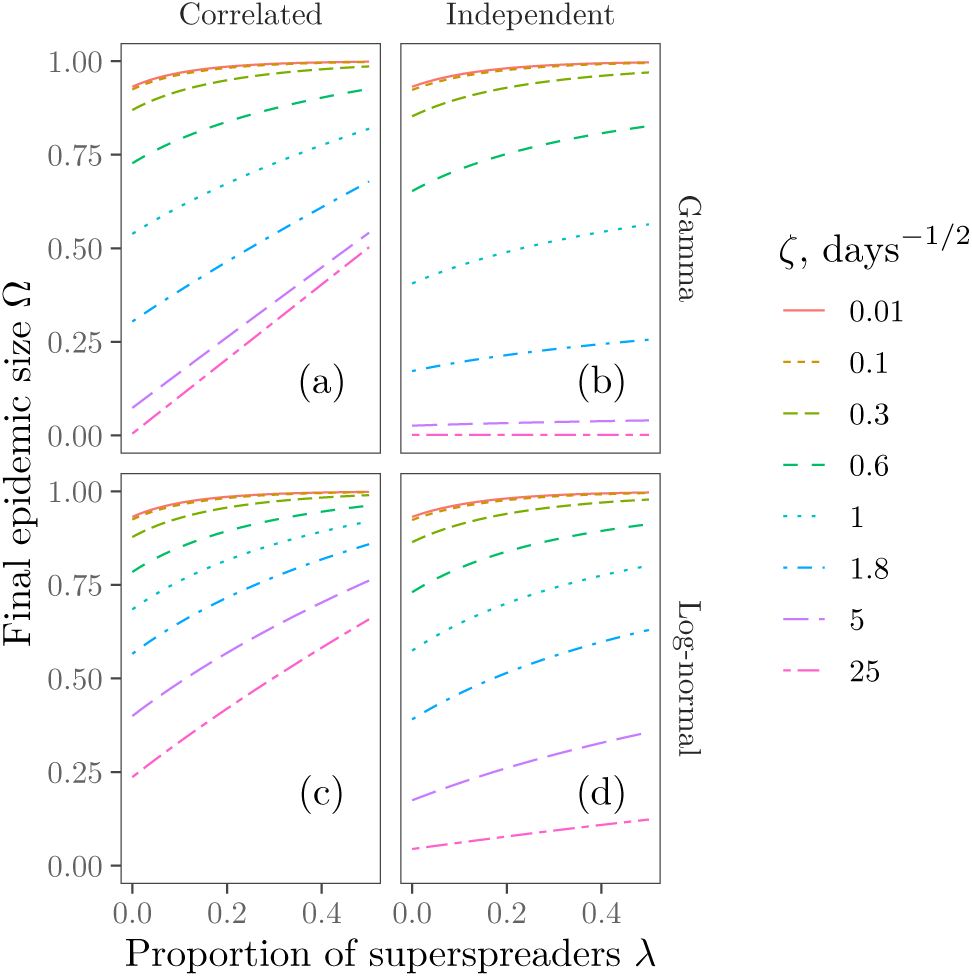
Final epidemic size for a mix of normal individuals (same distribution as on Figure 2) and superspreaders described by equation (33). The effect of superspreaders is at most linear in their proportion.

## V. DISCUSSION AND CONCLUSIONS

The aim of any idealized model is to provide insights about the “real world”. We believe our model provides several important insights beyond the assumptions involved in its derivation and treatment.

First, the variation in individual susceptibility and infectivity does matter. All examples studied in Section IV have the same average susceptibility and infectivity—but the outcomes greatly differ. Generally wider distribution lead to lower final epidemic size, and, in the case of correlated infectivity and susceptibility, a faster initial outbreak.

Second, the correlation between infectivity and susceptibility is important: the higher the correlation, the larger the epidemic size.

Third, the average and the width of infectivity and susceptibility are *not* enough to predict the outcome: the actual shape of the distribution matters too. The comparisons of log-normal and Gamma distributions in Figure 2, and of three different distributions having the same first and second moments in Figure 4, demonstrate this clearly.

This conclusion shows that a prediction of the epidemic’s spread is a hard task from the practical point of view. Indeed, one never knows the exact shape of the distribution, since it involves the measurement of individual infectivities and susceptibilities of a great many people. The sensitivity to the shape of the distribution beyond a couple of moments is bad news for precise predictions.

Having said this, we still need to answer the question of which features of the distribution are the most salient for predictions. There were a number of works stressing the importance of superspreaders: individuals or events with anomalously high potential for spreading (see the Introduction). Our model suggests a more nuanced view. On the one hand, because the susceptibility and infectiousness of individuals are correlated through how many people someone interacts with, increasing the number of superspreaders in a way that does not change the average infectivity or susceptibility will increase ℛ_0_ = 𝔼[*σs*]/*γ*, which greatly increases how fast the infection takes off and decreases the threshold for the outbreak and somewhat increases the final epidemic size. In the unrealistic case where we add pairs of one superspreader and one unusually careful person so that the variance increases and ℛ_0_ is unchanged, adding both these people will actually tend to decrease the final epidemic size. This can be seen in equations (21) and (22) where we have exponentials suppressing the contribution of individuals with anomalously high susceptibility (or high infectivity, if these parameters are correlated). This can also be seen in Appendix C and in Figure 4. The final result is determined by the average 𝔼[*σs*] *and* the distribution shape at low to moderate susceptibilities. It should be noted that, for wide distributions, the median *s* and mean *s* are quite different, and our conclusion concerns mean, rather than median, susceptibility.

Perhaps the following analogy may help to understand the meaning of this result. In comic books the outcome of a war is determined by a handful of superheroes and supervillains. In reality, by contrast, the final result is determined by the combined effort of many people at the lowest rungs of the military hierarchy: privates, petty and junior officers, and so forth. Our conclusion is that epidemic spread is like the “real war” rather than the “comic-book one”. The foregoing analysis has an essential implication for public health policy. While the prevention of superspreading is important (it changes the exponential growth rate ℛ_0_ = 𝔼[*σs*]/*γ* and drives down the averages in equations (21) and (22)), it is the mundane everyday efforts that matter most. Therefore, a way to localize the outbreak before mass vaccination becomes an option is to drive down the spread during many daily activities and perform rigorous tracing of everyday contacts. It is important to note that this discussion concerns superspreading *individuals*. The suppression of superspreading *events*, on the other hand, might be a powerful tool in preventing the spread.

In sum, we provide a simple, but efficient mathematical apparatus to calculate the epidemic dynamics for a population with variable infectivity and susceptibility, and cast it in a form suitable for numerical estimates. We hope this apparatus might turn out to be useful beyond the insights formulated in this paper.

Our model has some limitations that provide ample motivation for future work. One of the most important among them is the neglect of spatial inhomogeneity (the panmictic assumption). In reality each person has their own network of contacts, which is a source of inhomogeneity in infectivity [11]. The spread of infection is significantly influenced by the finite size of the individual’s contact network compared to the full population [7, 11, 43–46]. It would be very interesting to model the combination of spatial inhomogeneity *and* inhomogeneity in infectivity and susceptibility.

Lastly, operating in the current pandemic context, where long-lasting immunity has been observed, we have operated within the SIR paradigm, where a recovered person can never be infected again. If we allow for the reinfection of recovered individuals, such as in an SIRS or SIS model, we would expect superspreaders to have a much greater impact on the course of the epidemic. This is because their removal from the system at early times is now only temporary. Therefore, considering the possibility of reinfection will be a very important future application of our methods.

## Data Availability

Not applicable

## ACKNOWLEDGMENTS

The authors acknowledge the generous support of Chan Zuckerberg Initiative and Chan Zuckerberg Biohub. We are grateful to Dan Zigmond and Rob Phillips for their provocative questions and creative curiosity. We are also greatly indebted to interactions within the COVID-19 Discussion Group, including D. S. Fisher, M. Kamb, G. Le Treut and A. McGeever.

M. Rychnovsky is partially supported by I. Corwin’s NSF grant DMS:1811143 as well as the Fernholz Foundation’s “Summer Minerva Fellows” program. D.Y. acknowledges support by MINECO (Spain) through Grant No. PGC2018-094684-B-C21, partially funded by the European Regional Development Fund (FEDER).

## Appendix A Derivation of main equations

This Appendix is dedicated to the derivation of the main equation and the results of the general analysis in Section III.

First, we derive equation (13). Let us add equations (5) and (6) and use the definitions of *T* (*σ, s, γ, t*) to obtain

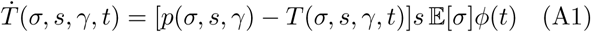

By inspection, we may verify that Eq.13 is a solution to this differential equation. We see that this solution satisfies the initial conditions

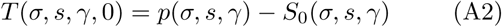

We now turn to the derivation of equation (14). First, we use equation (5) and the definitions of *T* (*σ, s, γ, t*) and *ϕ*(*t*) to write down

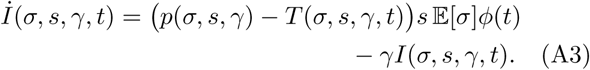

Substituting this expression into equation (A1), we arrive at

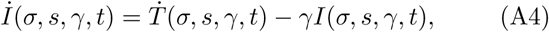

or, equivalently

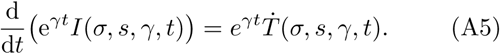

This differential equation admits a solution

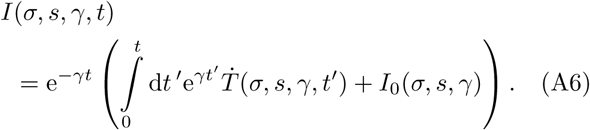

We now perform integration by parts in the above expression. In the second step and the second-to-last step, we will use our solution for *T* (*σ, s, γ, t*) from equation (13).

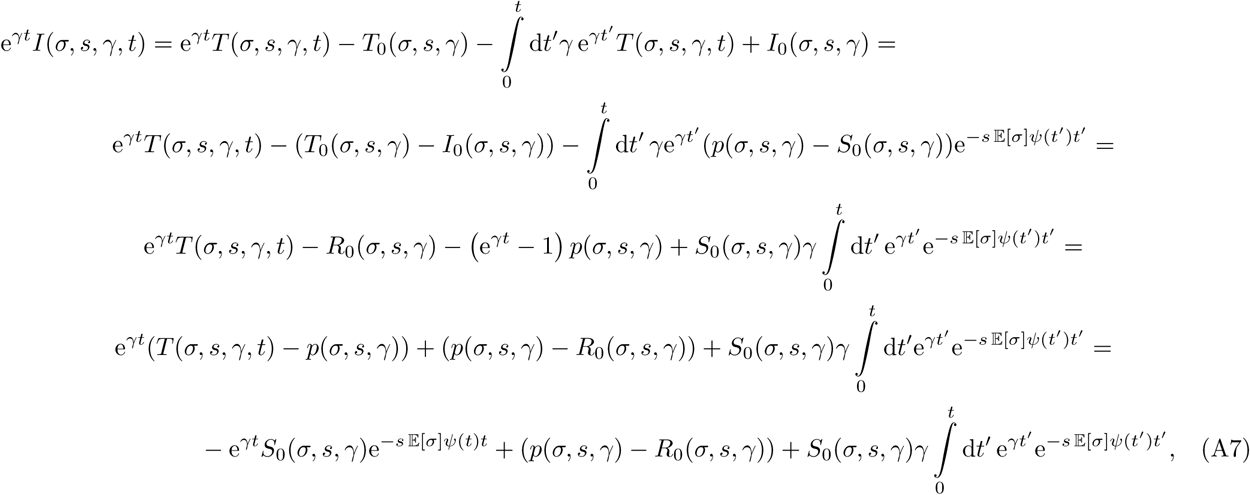

and therefore

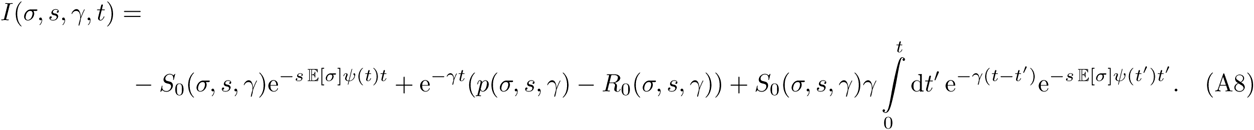

This final line matches Eq. (14).

Finally, we derive the equations of motion for *tψ*(*t*) as written in (15). We begin by substituting in our solution for *I*(*σ, s, γ, t*) into the definition of *ϕ*(*t*) in equation (10):

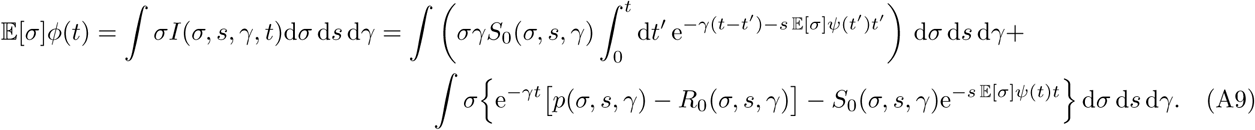

Noticing that *ϕ*(*t*) = d(*ψ*(*t*)*t*)/d*t*, we get

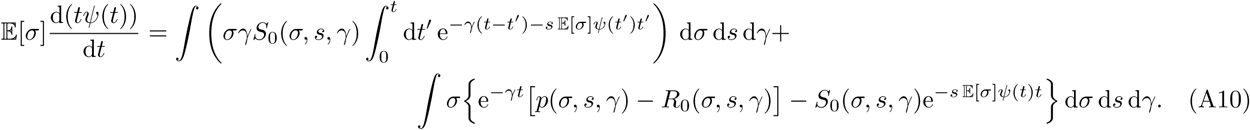

Integrating this equation by parts, we get

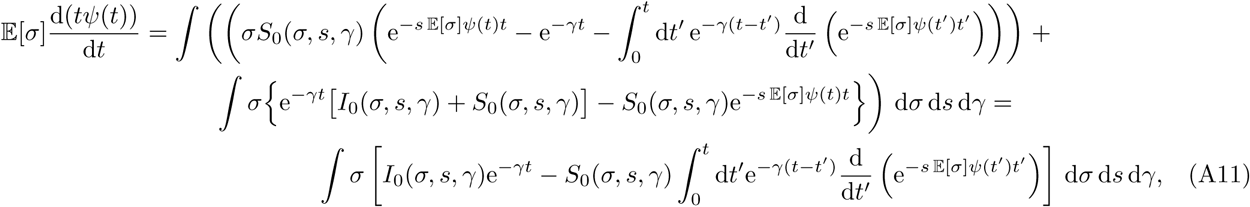

which matches equation (15).

Let us now derive equation (22) and propose an iterative algorithm for its numerical solution.

Assuming constant *γ* (equation (16)), we multiply both sides of equation (15) by e^*γt*^ and take a time derivative of both sides:

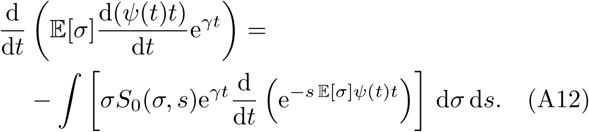

Taking the derivative of the left hand side, multiplying by e^−*γt*^ and integrating over time, we get

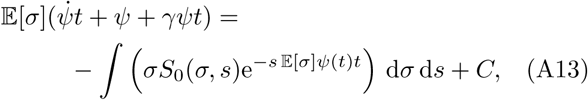

where *C* is a constant based on initial conditions. With the initial conditions (18), we get *C* = 𝔼 [*σ*]. As an aside, we may alternatively write equation (A13) as a first-order, time-independent equation using *ν*(*t*) = *ψ*(*t*)*t*.

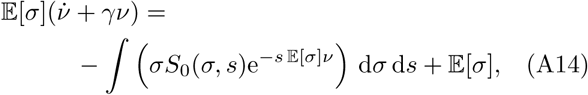

We rewrite as

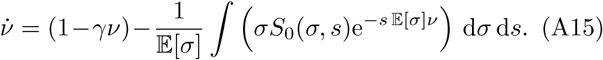

It is not hard to see that the right hand side is Lipshitz in *ν*, so the solution exists and is unique on ℝ_≥0_ by a standard application of the Picard-Lindelof theorem. In fact we have a bijection between solutions to the system (4), (5), (6) and solutions to (A15) given by

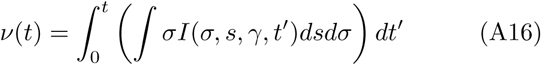

in one direction and by equations (13) and (14) in the other. Thus, existence and uniqueness of solutions to (A15) implies the existence and uniqueness of solutions to the system (4), (5), (6).

We already know that lim_*t*→∞_ *ψ*(*t*) = 0 (equation (12)). Suppose that 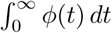 *dt* converges, and thus the following limit exists:

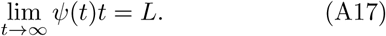

Then in the case *S*_0_(*σ, s*) = 1 − *ε* we obtain equation (22).

To justify the assumption (A17) we construct an algorithm to calculate *L* and prove it converges to a non-negative root of equation (22). We use the following iterations We will find the solution using the following iterations:

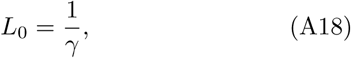

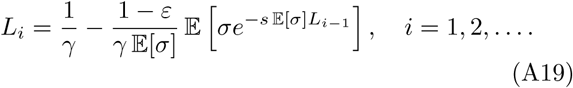

Below we will prove that the sequence *L*_*i*_ converges to the relevant root.

### Lemma 1.

*Suppose equation* (22) *has non-negative roots, and* 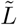 *is the largest root. Then the sequence L*_0_, *L*_1_, … *converges to* 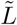.

*Proof*. We will prove that for all *i*

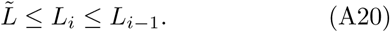

Then the sequence *L*_0_, *L*_1_, … is bounded and non-increasing, and therefore converges. The limit of this sequence is a root of equation (22), and due to inequality (A20) and the fact that 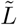 is the largest root, it converges to 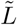.

First, note that from equations (22) and (A18) follows that 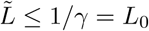.

For *i* = 1 we have from the iteration equation (A19) *L*_1_ ≤ *L*_0_ and, since 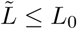,

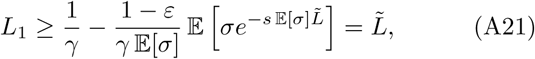

so inequality (A20) is true.

Suppose this inequality is true for *i* − 1, i.e.

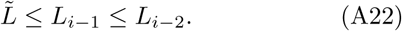

Then we will prove it for *i*. Indeed,

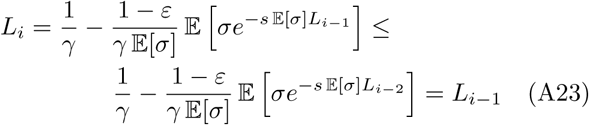

and

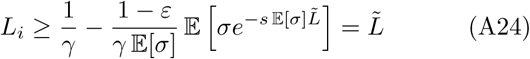

In other words if the inequality is true for *i* − 1, it is true for *i*, so it is true for all *i*. □

### Lemma 2.

*Equation* (22) *always has a non-negative root no smaller than ε/γ*.

*Proof*. Similarly to the proof of Lemma 1 we can prove the inequality

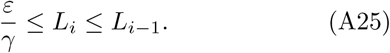

Indeed, for any *i* we can iteratively prove that

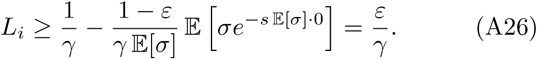

Therefore the sequence *L*_0_, *L*_1_, … converges to a number no smaller than *ε/γ*. This number is a root of equation (22), which, according to Lemma 1 is the largest root. □

The last lemma shows that the assumed behavior of *ψ*(*t*) at large *t* is indeed *ψ* ≈ *L/t*.

## Appendix B Short-time behavior and initial conditions

In this section we show that in a mixed population the parameter that determines whether an infection grows exponentially or dies out is

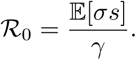

We also show that the long term behavior of the epidemic does not depend on the initial conditions.

At early time, when the proportion of the population infected, and the proportion of the population recovered are very small, equations (5) and (6) can be linearized as

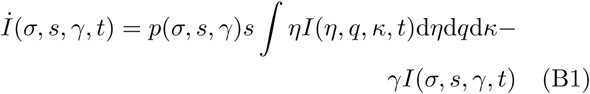

and

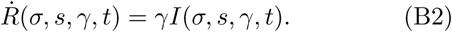

We consider the case where *γ* is fixed for the entire population, and the distribution 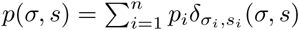 is a finite combination of delta functions. With the notation *I*_*i*_(*t*) = *I*(*σ*_*i*_, *s*_*i*_, *t*), equations (B1) and (B2) become a finite-dimensional system of equations

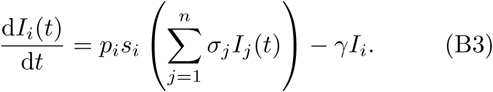

We rewrite this as

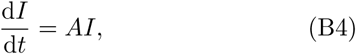

with *I* = *I*_1_ (*t*), … *I*_*n*_ ((*t*) ^*T*^ and *A*_*ij*_ = *p*_*i*_s_*i*_*σ*_*j*_ − *γ*𝟙_i=*j*_.

Let

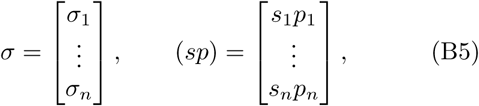

Let

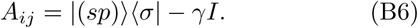

From this we see that the largest eigenvalue of *A* is 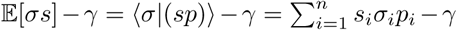 with the associated eigenvector |(*sp*) ⟩, and that all other eigenvectors are perpendicular to *σ* and have eigenvalue −*γ*.

Now a general distribution *p*(*σ, s*) can be approximated by a sum of point values, to conclude that the linear equations (B1) and (B2) have the largest eigenvalue

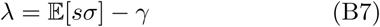

with corresponding eigenvector *I*(*σ, s*) = *sp*(*σ, s*) and all other eigenvalues negative.

If *p*(*σ, s*) is a compactly supported distribution we conclude that if a small enough proportion of the total population is infected at time zero, then until the proportion of the population that is susceptible drops appreciably below 1, we have

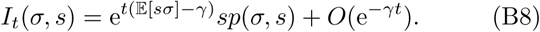

The quantity ℛ_0_ is also what epidemiologists measure when they measure the number of secondary infections produced by a typical infection in the very early stages of the epidemic. The key to understanding why this number is 𝔼 [*sσ*]/*γ* instead of 𝔼 [*s*] 𝔼 [*σ*]/*γ* comes from the word “typical.” Based on equation (B8), early in the epidemic the probability *q*(*σ, s*) that a person with infectivity *σ* and susceptibility *s* is infected is proportional to *sp*(*σ, s*), so

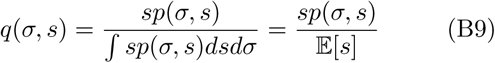

To find the number secondary infections per unit time this “typical infection” produces, we take this person’s infectivity and multiply by the average susceptibility in the population to get *σ*_typical_ 𝔼 [*s*]. Averaging *σ*_typical_ over the measure *q*(*σ, s*) gives

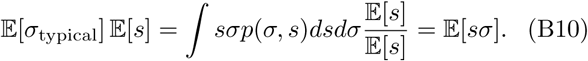

Multiplying by the typical recovery time 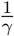 gives the expected number of secondary infections.

As with the usual SIR model, if ℛ_0_ *>* 1 the infection will spread and if ℛ_0_ *<* 1 the infection will die out. This allows us to see that the growth rate of an epidemic is highly dependent on how correlated *s* and *σ* are, with higher correlation leading to a higher growth rate. In a true population we expect a persons infectivity *σ* and susceptibility *s* to be highly correlated through factors like how many people someone interacts with. In particular superspreaders have an outsize effect on the early growth of the epidemic in the most realistic case where *s* and *σ* are highly correlated, because in this case ℛ_0_ grows like the second moment 𝔼 [*σ*^2^] of the infectivity rather than the first moment.

The second takeaway is that if the proportion of the population that is infected at time 0 is small enough, the solution for the system (5), (6) does not significantly depend on the details of the initial conditions. This can be seen by writing the initial profile of infected *I*_0_(*σ, s*) as a sum of eigenvectors for equations (B1) and (B2),

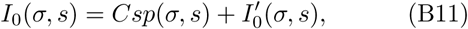

and comparing with (B8) to see that 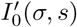 has minimal effect, and the long term solution is almost identical to the solution starting from initial condition

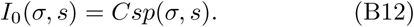

## Appendix C Worst-case distributions

In this section we discuss which distributions provide the highest possible epidemic size *Ω*_∞_ (the “worst-case scenarios”).

We prove two statements

i. *Variability is good*. If *s* and *σ* are independent, then the final epidemic size is less than or equal to the final epidemic size of the classical SIR model with *s*_0_ = 𝔼 [*s*], *σ*_0_ = 𝔼 [*σ*]. This conclusion agrees with the other studies of heterogeneity based on different assumptions and models [10, 11, 25, 29, 36, 37, 40].
ii. *Strong positive correlation is bad*. If the marginal distributions of *s* and *σ* are known, then the joint distribution *p*(*σ, s*) that maximizes the final epidemic size is given by the “percentile coupling”, where the *n*th most infectious person is also the *n*th most susceptible person.

Both these statements follow from the following lemma:

### Lemma 3.

*Let µ and ν be two possible joint distributions for* (*s, σ*). *Let* 𝔼^*µ*^ *and* 𝔼^*ν*^ *denote the expectation with respect to µ and ν respectively, and similarly for final epidemic sizes* 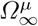*and* 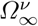. *If*

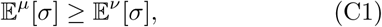

*and for all c >* 0,

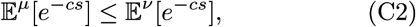

*and also*

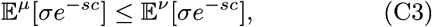

*then*

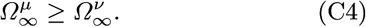

*Proof*. Using equations (C2) and (C1) together with equation (22) we see that for any *L >* 0,

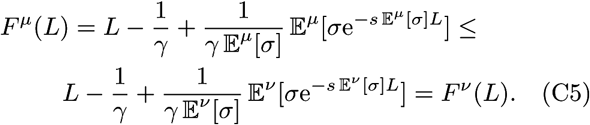

Let *L*^*µ*^ be the unique positive zero of *F*^*µ*^(*L*) if such a zero exists, and otherwise let *L*^*µ*^ = 0. Now *F*^*µ*^(0) = *F*^*ν*^ (0) = 0 and both are convex functions of *L*, which together with equation (C5) gives *L*^*µ*^ ≥ *L*^*ν*^.

Then from equations (C3) and (C1) we obtain

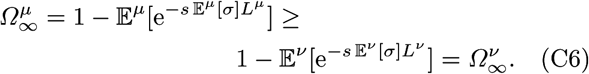

□

To prove (i) let us take a distribution *ν* with independent *σ* and *s*, and let *µ* = *δ*(*σ* − 𝔼^*ν*^ [*σ*])*δ*(*s* − 𝔼^*ν*^ [*s*]). We have 𝔼^*µ*^[*σ*] = 𝔼^*ν*^ [*σ*] by definition. From Jensen’s inequality [47, §1.7(iv)]

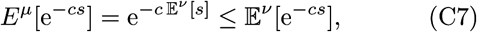

and from Jensen’s inequality and independence of *s* and *σ* under distribution *ν* we have

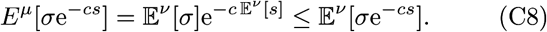

Thus the final epidemic size for our arbitrary distribution with independent *s* and *σ* is not greater than the final epidemic size of a delta mass with the same mean.

To prove (ii) let *ν* be an arbitrary measure with the correct marginal distributions, and let *µ* be the percentile coupling: the most susceptible person is the most infectious, the second most susceptible person is the second most infectious and so on. In particular if we sample twice from *µ* and obtain (*s*_1_, *σ*_1_) and (*s*_2_, *σ*_2_), then with probability 1, the statement *s*_1_ ≥ *s*_2_ implies *σ*_1_ ≥ *σ*_2_. This property implies that if *f* is an arbitrary decreasing function, and *g* is an arbitrary increasing function, then the percentile coupling is the coupling that minimizes the expectation E[*f* (*s*)*g*(*σ*)] for the given marginal distributions of *s* and *σ*. In particular this distribution minimizes *E*[*σ*e^−*sc*^] for all *c >* 0, so it satisfies equation (C3). It also has the same marginals as the other measure *ν*, thus inequalities (C1) and (C2) are satisfied. Thus for the given marginal distributions of *s* and *σ* the percentile coupling is the worst possible joint law in that it maximizes the final epidemic size of the infection.

To understand the meaning of statement i, consider the case of the population with the same infectivity, where some individuals have zero susceptibility, while all other individuals have the same large susceptibility *s*_1_. Let *f*_1_ be the fraction of these individuals. The epidemic size in this population does not exceed *f*_1_. To increase the variance of *s* while keeping the mean susceptibility constant, we must increase *s*_1_ and decrease *f*_1_, so large variance corresponds to lower epidemic size. Similarly one can consider a population with infectivity *σ* being either zero or a large value *σ*_0_ and show that when *σ*_0_ increases and the fraction of infectious individuals decreases the epidemic size also decreases.

Statement ii can be explained in the following way. Highly susceptible individuals become infected in the beginning of the epidemics, when the supply of susceptible individuals is high. If these highly susceptible individuals are also highly infectious, they can cause many secondary infections among the naïve population in this scenario, thus increasing the total size of the epidemics.

## Appendix D The effect of superspreaders

In this Appendix we discuss the effect of a superspreaders: a small subpopulation of people with anomalously high infectivity.

Consider the distribution of infectivity *σ* and susceptibility *s* as a sum of the “normal” distribution *p*_*n*_ and the superspreaders *p*_*s*_ with the latter having support at *σ > σ*_*s*_ with large *σ*_*s*_, as shown in equation (32).

The short term behavior is determined by the value of 𝔼 [*sσ*], which can be represented as

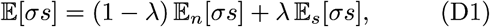

where subscripts *n* and *s* denote averaging with the distributions *p*_*n*_ and *p*_*s*_ correspondingly. This equation shows that (i) the only way superspreaders come into short term behavior is the renormalization of average *σs*, and (ii) their influence is linear in the proportion of super-spreaders *λ*.

Let us discuss the case where the number of super-spreaders is low enough, so the contribution of super-spreaders to the averages is small, i.e.

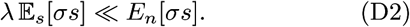

In this case the contribution of superspreaders into the short term dynamics is small according to equation (3). We are going to show that there is no anomalous contribution to the long term dynamics either.

We are looking into the final epidemic size, which is determined by equations (22) and (21).

First, consider the case where superspreaders have the same susceptibility distribution as the other individuals. In this case *s* and *σ* are independent, and our equations become

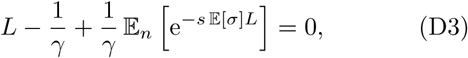

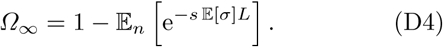

We see that in this case the only way superspreaders contribute is the changing of 𝔼 [*σ*].

Now consider the case where superspreaders have anomalous susceptibility *s*, and higher *σ* corresponds to higher *s*. Then the contribution of superspreaders is asymptotically small in both equations (22) and (21), i.e., again no worse than linear in the number of super-spreaders.

## Appendix E Numerically solvable equations

In this Appendix we will recast equation (15) into a set of differential equations suitable for numerical analysis.

With the constant *γ* assumption (16) and initial conditions (17) and (18), we can write down equation (15) as

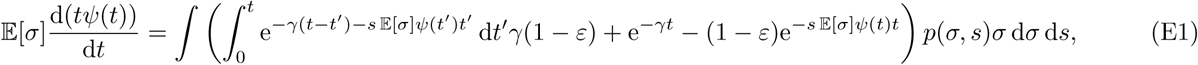

with

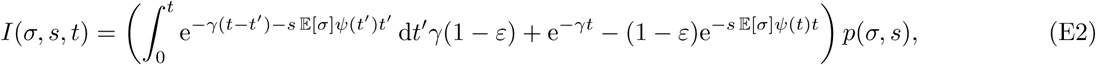

and

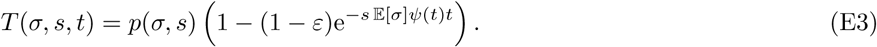

The initial condition is

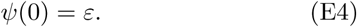

We introduce the function *ν*(*t*):

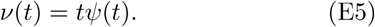

We multiply both parts of equation (E1) by e^*γt*^ and divide by 𝔼 [*σ*]:

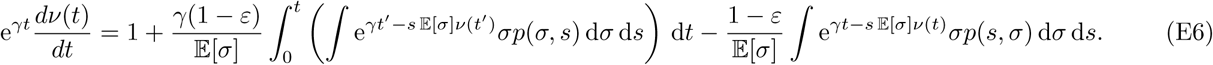

We differentiate this equation with respect to *t* and multiply by e^−*γt*^:

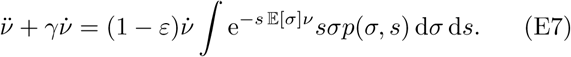

Let us introduce a new variable

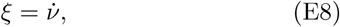

then we can write down equation (E7) as

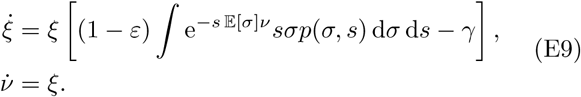

We need initial conditions for equations (E9). By definition (E5), *ν*(0) = 0. From equations (E8), (E5) and (E4) we get *ξ* (0) = *ψ*(0) = *ε*, so we can write initial conditions as

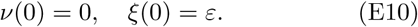

Equations (E9) with the initial conditions (E10) depend at any moment *t* on *ξ* (*t*) and *ν*(*t*) only, and therefore can be solved by any suitable method for differential equations.

## Appendix F Special distributions

For several important distributions we can provide analytical results. These results can be used for more sophisticated models, so we provide them below. We are particularly interested in the low-*γ* limit, where outbreaks are large and not easily controlled.

We discuss the completely correlated case when *σ*(*s*) is a monotonic function. Since we always can rescale them keeping *σs* constant, let us assume *σ* = *s*, so

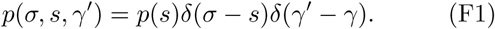

### 1. The Gamma distribution

Consider a Gamma distribution with fixed *γ* and *s* = *σ*, so *p*(*s*) in equation (F1) becomes

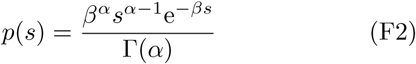

*α* and *β* being positive constants. First, let us calculate *L*, the root of equation (22). In our case we have

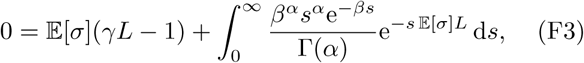

where 𝔼 [*σ*] = *α/β*. This gives for *L* the equation

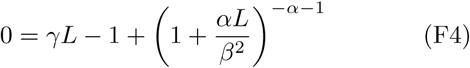

which can be easily solved numerically. The final epidemic size is given by equation 21, and may be written as

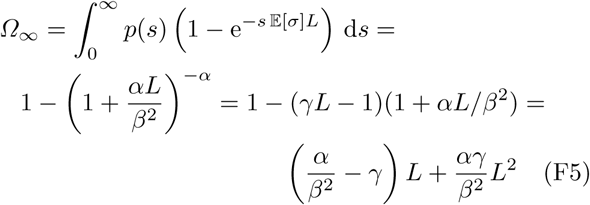

In the case of the exponential distribution (i.e., *α* = 1) equation (F5) becomes

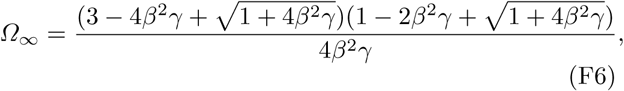

when ℛ_0_ *>* 1. We emphasize that (F5) and (F6) are exact formulas.

In the low *γ* limit we may approximate *L* by *L* = 1/*γ* − *f* (*γ*) (See equation (A19) and Lemma 1), where the second term can be written as

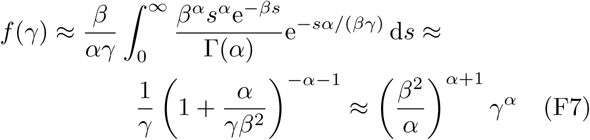

Since *α >* 0, *f* (*γ*) is well defined near *γ* = 0 and the approximation is well-controlled.

### 2. Low-recovery-rate limit for the log-normal distribution

Let us discuss a log-normal distribution with fixed *γ* and *s* = *σ*, where *p*(*s*) in equation (F1) becomes:

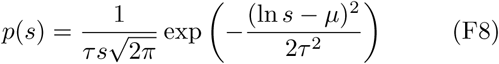

with the constants *τ >* 0 and *µ*. Note that due to equation (F1),

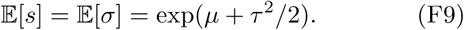

Using equations (A19) and (F9), we obtain the iterative equation for *ε* → 0:

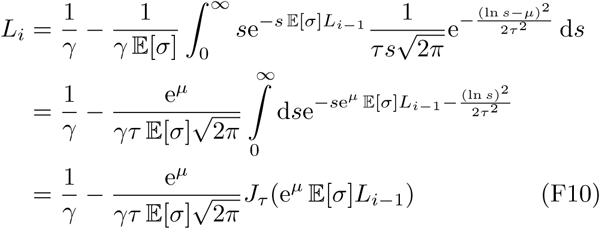

where we have defined

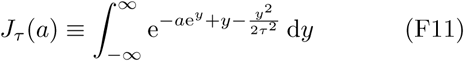

In principle, these equations are enough to construct an iterative solution for *L*. However, we may take this a step further for the low *γ* (large *L*) limit. In particular, if *L* is large, then so is each *L*_*i*_. For *a* ≡ e^*µ*^ 𝔼 [*s*]*L* ≫ 1, Eqn. (F11) can be evaluated by a standard saddle point approximation[48, 49]. Setting 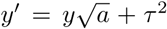 and expanding around 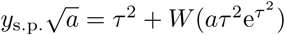 gives

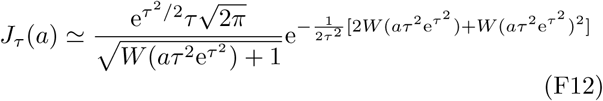

where 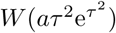 is the principal branch of the Lambert W-function, satisfying *W* (*ρ*) exp *W* (*ρ*) = *ρ*. This expression is valid up to a small correction of order *O*(*τ* ^2^/*W*) ∼*O*(*τ* ^2^/ ln(*a*)) ≪ 1.

Returning to our iterative solution for *L* in Eqn. (F11), we will now plug in the previous expression. Note that 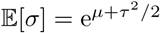

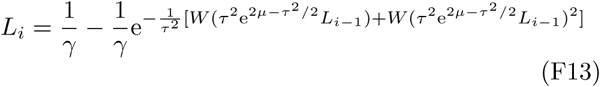

In particular,

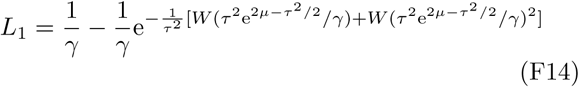

One may continue this iteration procedure to arbitrary precision.

## Appendix G SafeGraph Data

In this Appendix we describe the approach by Looi et al. [39] to transform the set of location pings into a dynamic network. In this network users are represented as nodes, and an edge (*u, v, t*) indicates that user *u* crossed paths with user *v* at time *t*. A path crossing is defined to occur when two users have pings which are separated by less than 50 meters and less than 5 minutes. It should be stressed that a path here is the same as a world line in relativity theory: it encompasses spatial and temporal dimensions, so the users cross paths if they are at the same place at the same time.

To ensure that users are represented accurately, various filters are applied; for example, excluding users with fewer than 500 pings or removing duplicate users, which could potentially occur if a single person carries multiple mobile devices. To compute the path crossings efficiently, the authors apply a sliding time window, and, within each time slice, use a k-d tree to identify all pairs of points within 50 meters of each other. We refer the reader to the original paper for details of the network-construction methodology. The constructed network captures 1 613 884 111 path crossings between 9 451 697 users across three evenly spaced months in 2017 (March, July, and November). The network provides an estimate of the true contact network, where each user’s number of contacts represents how many people they could possibly transmit the virus to or from. Thus, we can use each user’s degree in the path crossing network to estimate their susceptibility and infectivity.

Previous analyses of SafeGraph data have shown that it is representative of the US population, in that it does not systematically over-represent users from certain income levels, racial demographics, degrees of educational attainment, or geographic regions [50]. Recently, their mobility patterns have been instrumental in helping researchers study responses to the COVID-19 pandemic and to model the role of mobility in the spread of disease [7, 45, 51, 52]. Even so, there are caveats to the data that we use. Most notably, the path-crossing network covers three months in 2017, but individuals’ mobility patterns may have changed substantially following the onset of the pandemic. Furthermore, different types of noise may affect an individual’s number of observed crossings; for example, the frequency with which their phone pings. Filtering for only well-represented users can help to mitigate this issue.

